# Viral genetic determinants of prolonged respiratory syncytial virus infection among infants in a healthy term birth cohort

**DOI:** 10.1101/2022.06.22.22276752

**Authors:** Dylan Lawless, Christopher G. McKennan, Suman R. Das, Thomas Junier, Zhi Ming Xu, Larry J Anderson, Tebeb Gebretsadik, Meghan H. Shilts, Emma Larkin, Christian Rosas-Salazar, James D. Chappell, Jacques Fellay, Tina V. Hartert

## Abstract

1

**Background:** Respiratory syncytial virus (RSV) is associated with acute respiratory infection. We sought to identify RSV variants associated with prolonged infection.

**Methods:** Among healthy term infants we identified those with prolonged RSV infection and conducted 1) a human GWAS to test the dependence of infection risk on host genotype, 2) a viral GWAS for association with prolonged RSV infection using RSV whole-genome sequencing, 3) an analysis of all viral public sequences, 4) an assessment of immunological responses, and 5) a summary of all major functional data. Analyses were adjusted for viral/human population structure and host factors associated with infection risk.

**Results:** We identified p.E123K/D and p.P218T/S/L in G protein that were associated with prolonged infection (P*adj* = 0.01). We found no evidence of host genetic risk for infection. The RSV variant positions approximate sequences that could bind a putative viral receptor, heparan sulfate.

**Conclusions:** Using analysis of both viral and host genetics we identified a novel RSV variant associated with prolonged infection in healthy infants and no evidence supporting host genetic susceptibility to infection. As the capacity of RSV for chronicity and its viral reservoir are not defined, these findings are important for understanding the impact of RSV on chronic disease and endemicity.

**Key points:** Using a comprehensive computational analysis of viral and host genetics we identified a novel RSV variant associated with prolonged infection and no evidence supporting host genetic infection susceptibility, findings important to understanding RSV contribution to chronic disease and viral endemicity.

**Summary:** A comprehensive computational statistical analysis of both host and viral genetics provided compelling evidence for RSV viral persistence in healthy human infants, a finding of significant importance to understanding the impact of RSV on chronic disease and viral endemicity.

## 2 Introduction

Human orthopneumovirus, formerly known (and still referred to) as respiratory syncytial virus (RSV), results in significant global morbidity and mortality [1]. By the age of two to three years, nearly all children have been infected with RSV at least once [2]. RSV is a seasonal mucosal pathogen that primarily infects upper and lower respiratory tract epithelium, although it has been recovered from non-airway sources [3]–[8]. While RSV is mainly associated with acute respiratory infection, many RNA viruses can establish prolonged or persistent infection in some infected individuals [9]. Prolonged shedding of RSV, especially in young infants and following first infection, has been demonstrated, with longer average duration of viral shedding when polymerase chain reaction (PCR) is used to detect RSV [10]. While younger age and first infection are associated with protracted infection [2], [11], it is not known whether specific viral factors contribute to prolonged RSV infection in infants. This is important, as prolonged infection may contribute to enhanced transmission and developmental changes to the early life airway epithelium. Further, the reservoir of RSV infection is not understood, and it is possible that some RSV strains sustain a low level of ongoing viral circulation in the community until seasonal or other influences favor epidemic spread [12].

The objectives of this study were therefore to determine if there exist host or pathogen genetic risk alleles for RSV infection and to identify viral genetic variation associated with prolonged infection. These motivating questions are of fundamental interest in understanding viral and host genetic contributions that may underlie the development of chronic respiratory morbidity due to RSV, including asthma.

## 3 Methods

The protocol and informed consent documents were approved by the Institutional Review Board at Vanderbilt University Medical Center (#111299). One parent of each participant in the cohort study provided written informed consent for participation in this study. The informed consent document explained study procedures and use of data and biospecimens for future studies, including genetic studies.

Among healthy term infants in a cohort specifically designed to capture first RSV infection we identified those with prolonged RSV infection and conducted 1) a viral GWAS using RSV whole-genome sequencing to determine the relationship between viral genotypes and prolonged infant RSV infection, 2) a human GWAS to test the dependence of first year RSV infection risk on the genotype, 3) an analysis of all viral public sequence data, 4) an assessment of the local immunological RSV responses, and 5) a summary of all the major functional data for the identified viral variant. Full details of the methods are included in the Supplement, sections 8.1-8.13.

## 4 Results

### 4.1 Cohort characteristics

The INSPIRE cohort consisted of 1,949 enrolled infants among whom there were 2,093 in-person respiratory illness visits completed during winter virus season, November – March, of each year (Figure S1); the median (interquartile range [IQR]) number of in-person respiratory illness visits per infant during this surveillance window was 1 [1]. There were 344 RSV PCR-positive samples from 325 individuals which were sequenced. Prolonged infection was *a priori* defined as meeting criteria for acute respiratory infection with two or more RSV PCR positive nasal samples ≥ 15 days between testing, with improvement in symptoms between testing. Thus, these infections do not represent severe infections, but prolonged infections. These infections were on average less severe compared with term healthy infant infection among the entire cohort as measured by an ordinal respiratory severity score (Supplemental methods section 8.2) [12]. There were 19 infants who met the definition of prolonged infection with available viral sequencing used to confirm clonality of original and subsequent virus detections. The mean RSV Ct value of first infections was 25.9 ± 7.1, and second detection was 31.6 ± 5.4. All samples were analyzed together and raw values are reported without normalizing Ct to housekeeper genes. The mean number of days between detections was 29 ± 21 days (Figure S2). Table 1 lists the cohort characteristics of infants with prolonged RSV infection compared with other RSV infection and the entire cohort.

**Table 1:** Characteristics of infants with prolonged RSV infection compared with other RSV infection and the entire cohort. Prolonged infection is defined as RSV PCR-positive samples with ≥15 days between testing and meeting criteria for acute respiratory infection. *Presence of sibling or another child ≤ 6 years of age at home.

|  | **Prolonged RSV**<br>**infection**<br>**N = 19** | **RSV infection**<br>**N=342** | **Total**<br>**N = 1949** |
| --- | --- | --- | --- |
| Age in months at first RSV illness (median, IQR) | 6 (4, 6) | 4 (2,5) | NA |
| Illness respiratory severity score | 2.0 (1.2, 3.0) | 3.0 (2.0, 4.0) | NA |
| RSV season |  |  |  |
| 2012-13 | 68% | 54% | 44% |
| 2013-14 | 32% | 46% | 56% |
| Self-reported race |  |  |  |
| Non-Hispanic Black | 37% | 13% | 18% |
| Non-Hispanic White | 63% | 69% | 65% |
| Hispanic | 0% | 10% | 9% |
| Multi race/ethnicity/other | 0% | 8% | 8% |
| Female sex | 53% | 44% | 48% |
| Second-hand smoke exposure | 21% | 23% | 47% |
| Health insurance |  |  |  |
| Medicaid | 68% | 48% | 54% |
| Private | 32% | 51% | 45% |
| None/unknown | 0% | 1% | 1% |
| Daycare and/or siblings | 84% | 78% | 66% |

### 4.2 Host genetic analyses

We explored whether RSV infection in infancy is a natural assignment (quasi-random) event and, unlike severity of early-life RSV infection [13], occurs independently of host genetics. For the candidate SNP analysis, we considered childhood asthma- and RSV LRTI-associated SNPs identified in Pividori et al. [14]; Janssen et al. [15]; Pasanen et al. [16] (methods). Associations between genotype at the resulting 54 SNPs (50 childhood asthma- and 4 RSV LRTI-associated SNPs) and RSV infection in infancy in our data are given in Figure 1. The data are consistent with little to no effect of genotype at these SNPs on RSV infection in infancy.

**Figure 1:**
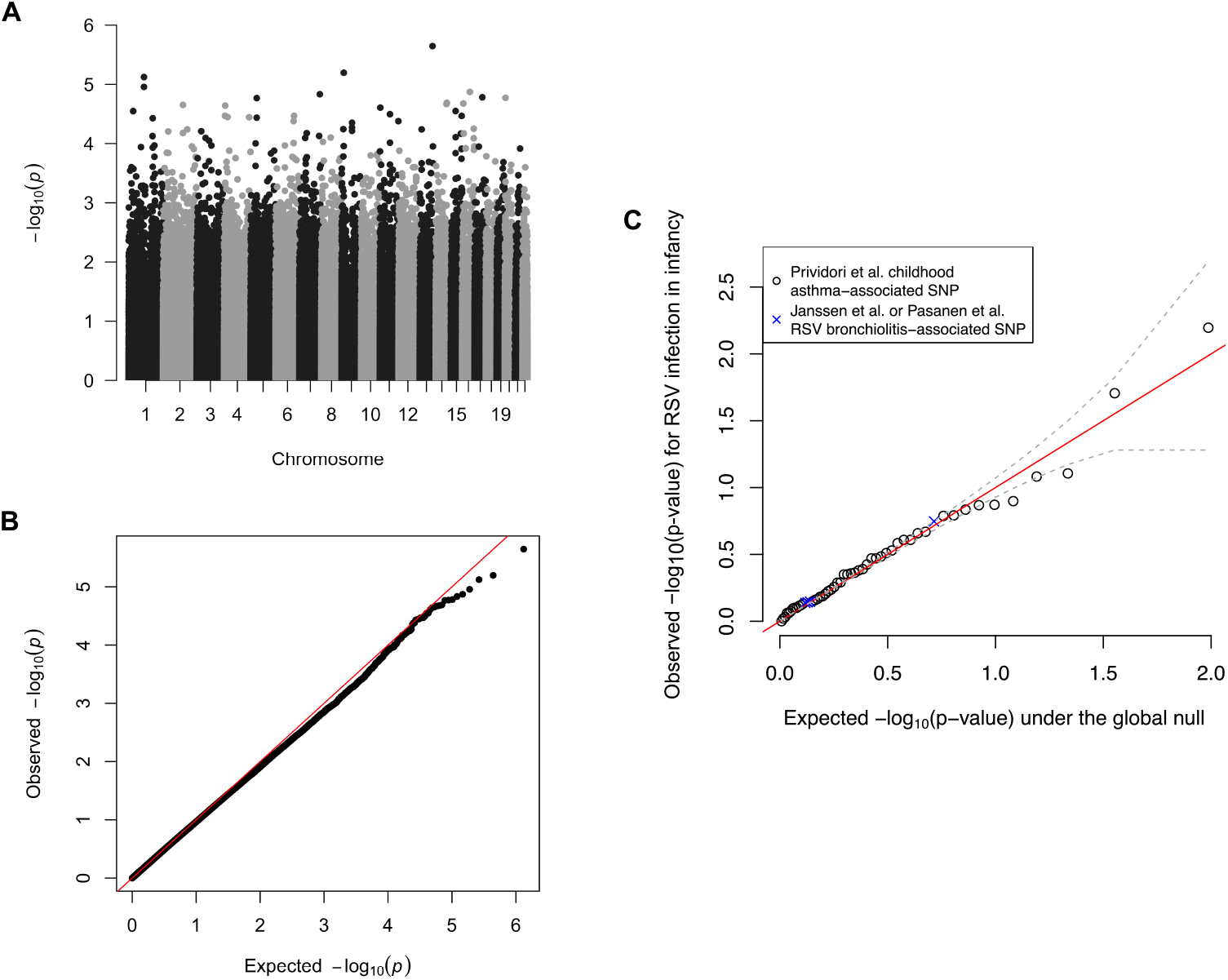
Genetic analyses of RSV infection in infancy. (A) The Manhattan plot shows no genome-wide significant associations (p value threshold of 5e-8). (B) The Q-Q plot demonstrates that the observed p values are congruent with those expected under the null hypothesis that RSV infection in infancy is independent of host genotype. (C) The association between the 54 selected childhood asthma- or RSV LRTI-associated SNPs and RSV infection in infancy in our data. The identity line is shown in red, and the dashed grey lines are ±1 standard deviation around the expected *-log10*(p value). RSV: respiratory syncytial virus; SNP: single nucleotide polymorphism.

We further investigated the possibility that the analysis was underpowered to identify associations with these SNPs by pooling information across SNPs to estimate the average genetic effect size [17]. We estimated the narrow-sense heritability of RSV infection during infancy on the latent liability scale (h^2^_l_), which, if > 0, would indicate an accumulation of small genetic effects. We estimated h^2^_l_ to be exactly 0, suggesting that, if present, infant RSV infection-related genetic signals are both small and sparse (Supplemental sec 8.6).

### 4.3 Population structure

A summary of protein coding genes in RSV is illustrated in Figure 2 A. Our analysis focused on F and G protein. The phylogenetic tree based on multiple sequence alignment (MSA) of G protein amino acid sequences is shown in Figure 2 B. One obvious feature causing a separation in genetic diversity is G protein partial gene duplication, which has emerged in recent years within RSV-A strains [18]. RSV-B strains with a homologous duplication have existed for two decades, although the selection process leading to emergence and clinical implications have not been entirely defined.

**Figure 2:**
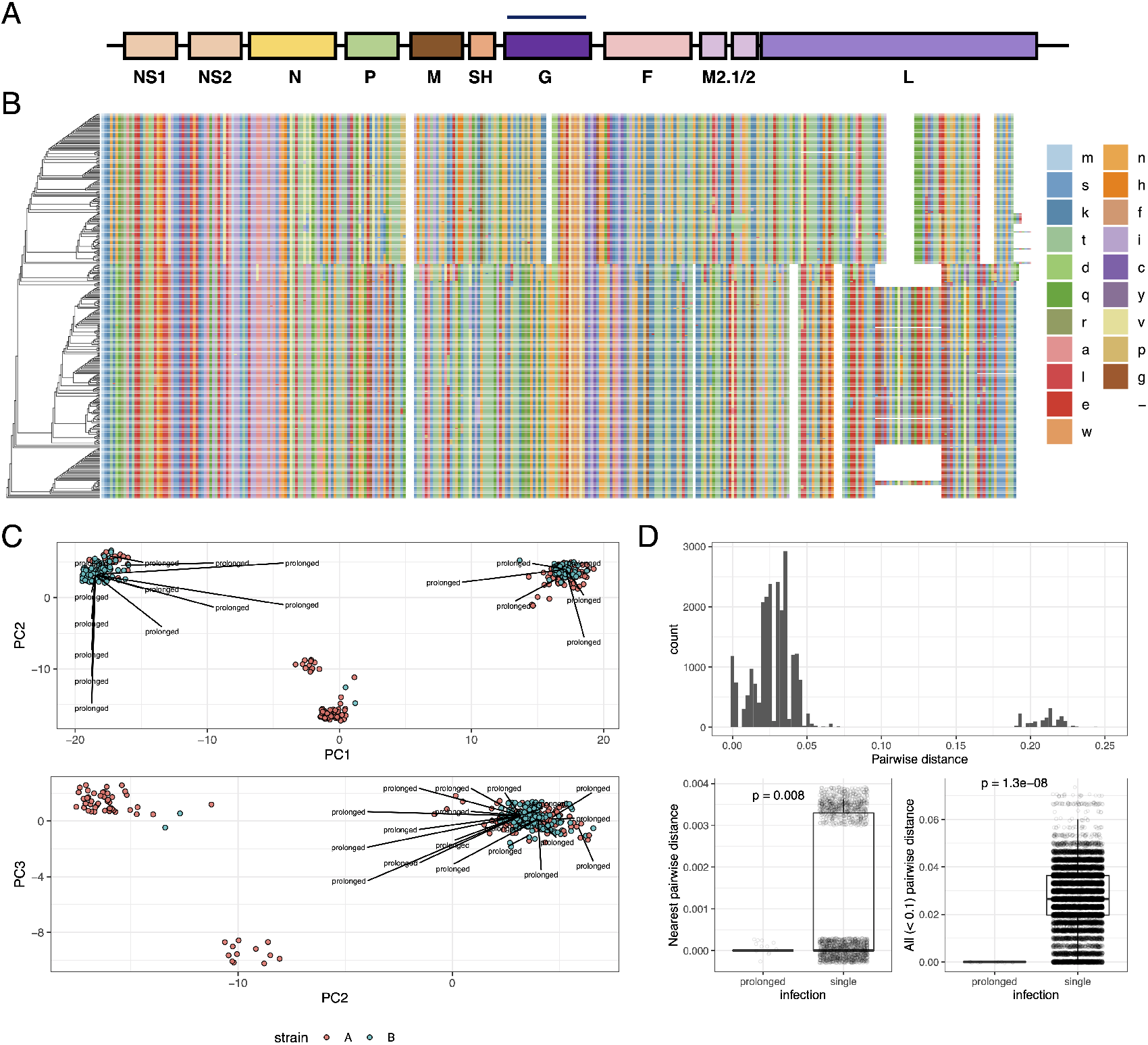
Viral population structure. (A) Linear map of the RSV genome. (B) Phylogenetic tree based on multiple sequence alignment MSA of G protein amino acid sequences. Color; amino acids. (C) Principal component (PC) analysis. PCs1-3 with labels indicating prolonged infections from different phylogenetic clades. (D) Panel [i] summarises every pairwise genetic distance between every viral sequence. Genetic invariance in prolonged infections separated by at least 15 days compared to other genetic variation within the most closely related sequences (panel [ii]) and within all possible closely related pairs (panel [iii]). VE (variance explained). Jitter applied for visualisation.

PCA was used for reducing the dimensionality of sequence data, where PC1 accounted for 95.19% of cumulative variance, and variance attributed to other PCs was roughly uniformly distributed (Figure 2 C). We observed prolonged infections by viruses from different phylogenetic clades, rather than one specific clade (Figure 2 C), indicating that these results are not confounded by latent clade membership.

### 4.4 Genetic invariance of prolonged infection

Figure 2 D panel [i] summarizes every pairwise genetic distance between every viral sequence, where small distances indicate pairs with closely related sequences. Panels [ii] and [iii], which summarize the difference in sequence similarity distributions between viruses from the same host and different hosts, show that RSV sequences corresponding to initial and subsequent viral detections are nearly identical. These results support the conclusion that such cases are prolonged (i.e., failure to clear) infections rather than new infections.

### 4.5 Variants in G glycoprotein significantly associated with prolonged infection

Variants at the amino acid level were assessed for their association with prolonged infection. The model consisted of (A) the binary response (prolonged infection Yes/No), and (B) predictors; (1) viral genotype (REF/ALT amino acid), (2) viral PCs 1-5, (3) host sex, and host features that have been previously demonstrated as significantly associated with infection; (4) self-reported race/ethnicity, (5) child-care attendance, or living with another child ≤ 6 years of age at home [19]. A significant genetic association was identified between prolonged infection and the lead variant after Bonferroni correction for multiple testing (threshold for number independent variants < 0.05/23 = 0.002), as shown in Figure 3 A, p value = 0.0006.

**Figure 3:**
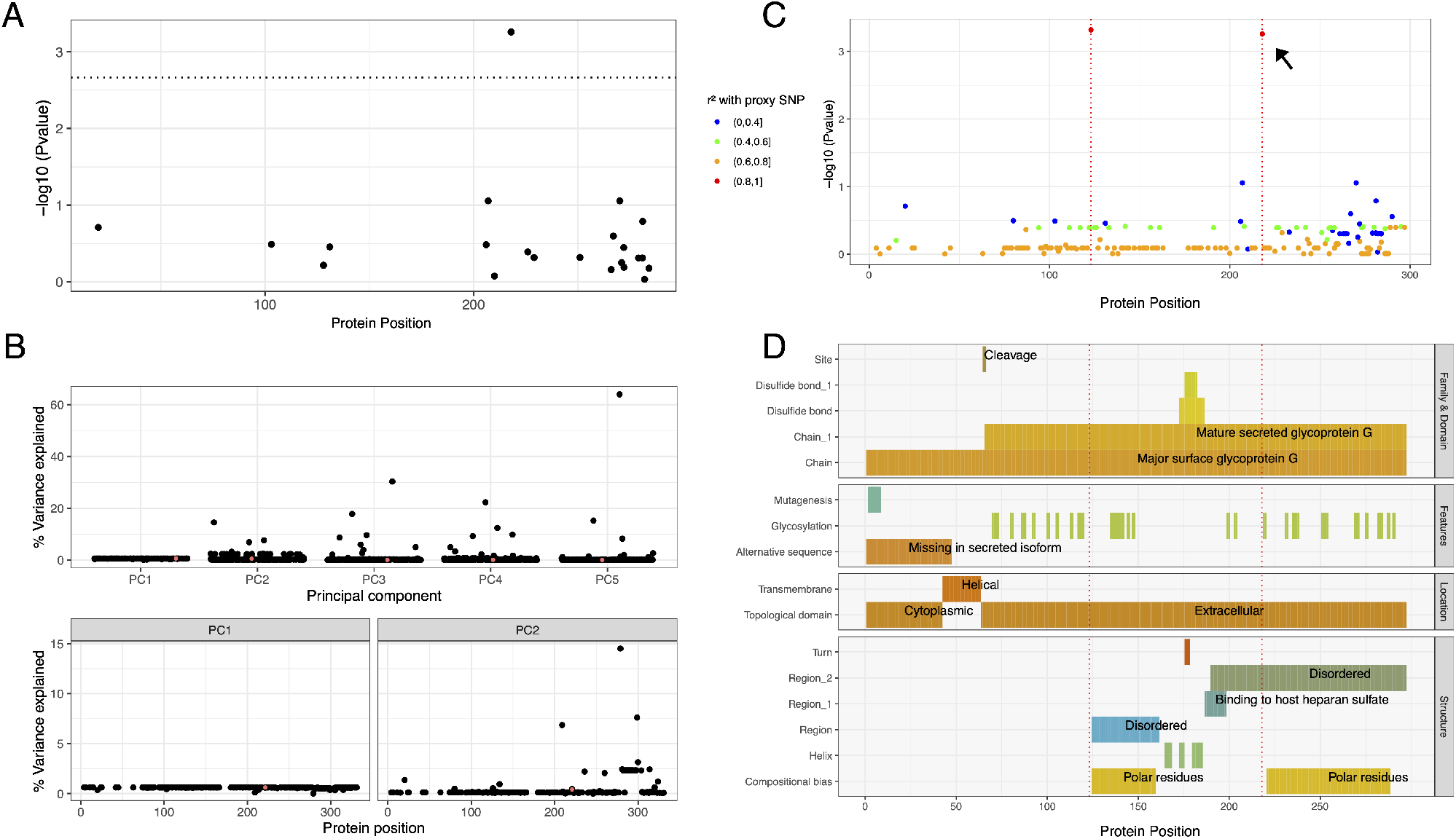
Viral genetic association with prolonged infection. (A) Amino acid association with prolonged infection after multiple testing correction (significant threshold shown by dotted line). (B) Variance explained (VE) within cohort. The effect of each variant on cohort structure is shown for PCs1-2. The small % VE for a significantly associated lead variant supports a true positive. (C) Variants in strong correlation were clumped for association testing using proxies for r^2^ ≥ 0.8. One significant association was identified (shown in A); the r^2^ values for all other variants show a single highly correlated variant with the lead proxy (red), identifying p.E123K/D and p.P218T/S/L. (D) Evidence for biological interpretation for every amino acid position is summarised. Dotted red lines indicate the positions at p.123, p.218.

To determine whether this association was simply due to population stratification between strains A and B, a subset analysis was performed using independently assessed clinical laboratory strain labels for A and B. The same direction of effect indicated that the association was not a false positive, although in this significantly smaller sub-analysis the result was not significant.

To assess the possibility of a false positive association due to population structure within our cohort, we assessed the magnitude of variance explained (VE) at every amino acid position. Figure 3 B (panel [i]) shows the variance explained by each amino acid in PCs1-5. The cumulative proportion of variance for PCs 1-5 was 99.5% (PC1 = 95%, PC2 = 3%). The values are illustrated according to protein position in panels [ii-iii]. The lead association variant had 0.603% VE for PC1 and 0.458% VE for PC2, a negligible effect that precludes spurious association by allele frequency between populations.

After identifying a significant viral genetic association with prolonged infection, we quantified the correlation of variants with the lead variant. Clumping was performed by ranking based on MAF and with a cut-off threshold of r^2^ ≥ 0.8 (Supplemental Figure S3). The association model was repeated for all variants, defining protein p.E123K/D and p.P218T/S/L as candidate causal variants associated with prolonged infection as shown in Figure 3 C. No other variants were correlated with this outcome.

To determine whether p.E123K/D and p.P218T/S/L variant genotypes are novel and potentially influence viral fitness, we searched public worldwide viral data (1956 onward), yielding a total of 1084 G protein sequences. The variants were present at a low and stable frequency, without obvious temporal enrichment (Supplemental Figure S4). Thus, while historical data reveal no positive selective advantage attached to p.E123K/D and p.P218T/S/L, longstanding circulation and linkage in prolonged RSV infection suggest that these polymorphisms are present in the viral inoculum and do not arise through recurrent mutational events.

Due to multiple testing correction according to our analysis plan, an association also originally identified in F protein was rejected and therefore omitted from further discussion. For posterity, the variant position was p.N116S (relative to strain A GenBank: AMN91253.1) (P = 0.0026; within F protein P_adj_ = 0.021, combined F and G P_adj_ = 0.082).

### 4.6 Functional interpretation

Cell-attachment proteins of paramyxoviruses (G protein in RSV) span the viral envelope and form spike-like projections from the virion surface. RSV G protein is a type II integral membrane protein consisting of 298 amino acid residues comprising N-terminal cytoplasmic (p.1-43), transmembrane helical (p.43-63), and extracellular (p.64-298) domains (Figure 3 D). RSV G protein ectodomain also exists in a soluble secreted form, p.66 – 298, which functions in immune evasion [20]–[22]. G protein interacts with the small hydrophobic (SH) protein [23] and, via the N-terminus, with matrix (M) [24] protein. It has also been reported to form homo-oligomers [25]. The variant amino acid positions associated with prolonged infection reside in a portion of the G protein ectodomain of unassigned specific function and linearly non-contiguous with sequences that bind cell-surface heparan sulfate, which likely promotes RSV cell-attachment (p.187-198) [20]–[22]. However, recent studies indicate that heparan sulfate is not present on the cell surface in the natural environment [1], [26], [27]. In addition, these positions do not contribute to known neutralization epitopes on G protein. Information available in PDB was insufficient to infer effects of p.E123K/D and p.P218T/S/L on local or regional protein structure. The potential effect on glycosylation is indeterminate. Lee et al [28] and Collarini et al [29] report on broadly neutralising antibodies which bind to p.164-176 (conserved sequences shared by both RSV A and B subtypes) and p.190-204, as well as CD4 epitopes within the latter region. Figure 3 D illustrates the position of the variants of interest relative to summarised known functional features.

## 5 Discussion

In this study of term healthy infants, we found no evidence of host genetic susceptibility to RSV infection during infancy. This allowed our analysis to focus on elucidation of viral drivers of prolonged infection. A significant viral genetic association in the RSV G protein, p.E123K/D and p.P218T/S/L, with prolonged infant RSV infection was identified. These variants were not associated with severe disease, and public data reveal their consistent presence at low frequencies over the past 30 years, without evidence of enrichment by positive selective pressure over time. The two variants we identified in G are correlated with non-random association analogous to LD in the human diploid genome and therefore not likely random mutations, but instead co-inherited in the infecting inoculum. This suggests an evolutionary benefit and raises the question of why such variants have maintained a stable but low frequency in the human population for at least four decades. These strains are a potential reservoir, emerging seasonally in response to immune, environmental, or other forces. Alternatively, the polymorphisms might recurrently arise de novo during infection of some individuals but are poorly transmissible because of suboptimal fitness. The possibility of viral mutational immune escape has been reported for infants who struggle to control primary RSV infections, allowing for prolonged viral replication and not previously described viral rebound [30].

The RSV variants associated with prolonged infection in our cohort, G p.E123K/D and p.P218T/S/L, lie in the surface region, and there are no known mechanistic features that directly overlap, although it is possible that variant positions approximate sequences that bind a putative viral receptor, heparan sulfate [21], in the G protein three-dimensional structure. While immortalised cell lines abundantly express surface glycosaminoglycans including heparan sulfate, recent work reports that RSV infects the apical aspect of ciliated respiratory epithelial cells, which lack detectable surface heparan sulfate [26], [27], [33]. G protein amino acid positions 123 and 218 are not part of known antibody neutralization epitopes or CD8+ cytotoxic T-cell epitopes (Figure 3 D). However, p.164-176 and p.190-204 are bound by broadly neutralising antibodies and CD4 epitopes are known within the region [28], [29]. Treatment and prophylaxis may be gained from the use of antibodies which target F and G proteins. In addition to heparan sulfate, interactions between G protein and CX3CR1, the receptor for the CX3C chemokine fractalkine, have been reported to modulate the immune response and facilitate infection [20]–[22] [27], [31], [32]. Furthermore, the mature secreted isoform of G protein (p.66-298) is thought to facilitate viral antibody evasion by acting as an antigen decoy and modifying the activity of leukocytes bearing Fc-gamma receptors [34]. Our findings raise the interesting prospect that G protein variants associated with prolonged infection alter a key interaction at the immune interface between pathogen and host.

Although this study was not designed to define mechanisms underlying the association of G protein variants with prolonged infection, these sequence changes might dampen antiviral immune responses and thereby delay viral clearance. It is possible that strains harbouring G protein p.E123K/D and p.P218T/S/L variants are cleared more slowly and foster an immune environment of low-level chronic stimulation or exhaustion. We previously demonstrated that infants infected with RSV in their first year of life have dampened subsequent antiviral immune responses in early childhood [36] as well as changes in airway epithelial cell metabolism [37]. Altered immune responses are expected in extended infections by G protein variant strains [35], and we observed differences in the acute antiviral response between subjects with resolved and prolonged infection, specifically increased levels of types 1 and 2 IFN in nasal secretions; however, we could not make causal inference about variant sequences because of confounding by co-linearity of these polymorphisms with RSV antigenic group.

While this study has a number of significant strengths, including one of few population-based surveillance studies of first RSV infections during infancy among term healthy infants, our findings are also subject to some limitations. First, this study was not designed with the primary intention to examine infection duration, and additional sampling following initial RSV infection was triggered by a repeat acute respiratory illness. Asymptomatic prolonged infections would therefore not have been captured. Second, our study cohort was small, necessitating focus on viral surface glycoproteins, F and G, due to their variability and importance in host immunity. A larger cohort with serial sampling would be required to diminish the impact of co-linearity of viral genotypes with antigenic groups and to perform informative viral whole genome analysis. Genome-wide information might elucidate other determinants of prolonged infection or pathogen fitness that mediate and/or modulate effects of phenotype-driving variations. Third, again due to small sample size, we could only investigate host genetic risk for infection, not prolonged infection. While we have not specifically assessed subjects for rare monogenic variants that may underlie immunodeficiency, our enrolment criteria included only infants who were term and otherwise healthy.

While we performed an interaction analysis for the outcome of host asthma, host genetics, and pathogen genetics and found no significant interaction, our sample size is unlikely sufficient to exclude such an interaction. Lastly, while we do not expect a role for immune memory in these first-in-life RSV infections, we cannot exclude modulatory effects of maternal antibody, which we did not measure. Despite these limitations, the results are novel and represent an in-depth comprehensive computational statistical analysis of both host and viral genetics providing compelling evidence for RSV viral strain persistence in healthy human infants, a finding of significant importance to understanding the impact of RSV on chronic disease and viral endemicity.

In summary, we identified a novel RSV viral variant associated with prolonged infection in healthy infants, but no evidence of host genetic susceptibility to infant RSV infection. Understanding host and viral mechanisms that contribute to prolonged infection will be important in crafting strategies to control the short- and long-term impact of RSV infection. The identification of RSV variants associated with prolonged infection might also improve vaccine design, particularly if these variants stimulate robust immunity or, in contrast, escape the immune response or induce immunopathologic conditions. The growing availability of large genomic and functional data sources provides opportunities for advancing our understanding of the pathogenesis of infant RSV infection, defining the contribution of viral genetic variants to acute and chronic disease, and informing the development of effective vaccines. As neither the capacity of RSV for prolonged infection in immunocompetent hosts nor a viral reservoir has been delineated, these results are of fundamental interest in understanding viral and host genetic contributions that may promote prolonged infection and influence development of chronic respiratory morbidity.

## 6 Links

### 6.1 Software

R v4.1.0 was used for data preparation and analysis http://www.r-project.org.

R package caret was used for analysis: genetic correlations.

R package dplyr was used for data curation.

R package factoextra was used for analysis: PCA, and to visualise eigenvalues and variance.

R package ggplot2 was used for data visualisation.

R package MASS was used to analysis: logistic regression model data.

R package stats was used for analysis: including glm for logistic regressions.

R package stringr was used for data curation.

R package tidyr was used for data curation.

asn2fsa https://www.huge-man-linux.net/man1/asn2fsa.html

clc novo assemble qiagenbioinformatics.com

Clustal Omega https://www.ebi.ac.uk/Tools/msa/clustalo/

dbNSFP (database) http://database.liulab.science/dbNSFP [38]

GCTA https://cnsgenomics.com/software/gcta/ [39]

GenBank https://www.ncbi.nlm.nih.gov/genbank/IQ-Tree https://www.iqtree.org/ [40]

KING https://people.virginia.edu/~wc9c/KING/ [41]

MAFFT https://mafft.cbrc.jp/alignment/software/ [42]

NextAlign https://github.com/nextstrain/nextclade

PLINK http://zzz.bwh.harvard.edu/plink/ [43]

Tbl2asn https://www.ncbi.nlm.nih.gov/genbank/tbl2asn2/

Viral Genome ORF Reader, VIGOR 3.0 https://sourceforge.net/projects/jcvi-vigor/files/

RCSB PDB https://www.rcsb.org

UniProt https://www.uniprot.org

### 6.2 Data sources

Dataset https://www.ncbi.nlm.nih.gov/bioproject/267583.

Dataset https://www.ncbi.nlm.nih.gov/bioproject/225816.

J. Craig Venter Institute https://www.jcvi.org.

GenBank:NC 001989 Bovine orthopneumovirus, complete genome

https://www.ncbi.nlm.nih.gov/nuccore/NC_001989.

Reference data https://www.ncbi.nlm.nih.gov/gene/?term=1489824. G attachment glycoprotein [Human orthopneumovirus]; ID: 1489824; Location: NC 001781.1 (4675..5600); Aliases: HRSVgp07.

Reference data https://www.ncbi.nlm.nih.gov/gene/?term=37607642. G attachment glycoprotein [Human orthopneumovirus]; ID: 37607642; Location: NC 038235.1 (4673..5595); Aliases: DZD21 gp07.

Reference data for all public NCBI Virus https://www.ncbi.nlm.nih.gov/labs/virus/vssi/ for species: Human orthopneumovirus; genus: Orthopneumovirus; family: Pneumoviridae.

Reference data https://www.ncbi.nlm.nih.gov/labs/virus/vssi/#/virus?SeqType_s=Nucleotide&VirusLineage_ss=Human%20orthopneumovirus,%20taxid:11250 contains sequence data for Virus Lineage ss=Human orthopneumovirus, taxid:11250 nucleotide: 26’965, protein: 53’804, RefSeq Genomes: 2.

Reference https://www.ncbi.nlm.nih.gov/protein/NP_056862.1 GCF 002815475.1 (release 2018-08-19) Nucleotide Accessions: NC 038235.1, protein: Y 009518856.1

Reference https://www.ncbi.nlm.nih.gov/protein/YP_009518856.1 GCF 000855545.1 (release 2015-02-12) Nucleotide Accessions: NC 001781.1, protein: NP 056862.1 (strain B1).

## Data Availability

All data produced in the present study are available upon reasonable request to the authors

https://github.com/DylanLawless/inspire2022lawless.github.io

## Abbreviations

ALT: (alternative)
CI: (confidence interval)
GWAS: (genome-wide association study)
G: (glycoprotein)
H: (hemagglutinin)
HN: (hemagglutinin-neuraminidase)
IFN: (interferon)
IQR: (interquartile range)
INSPIRE: (The INfant Susceptibility to Pulmonary Infections and Asthma Following RSV Exposure)
LD: (linkage disequilibrium)
LRTI: (lower respiratory tract infection)
MAF: (minor allele frequency)
MFI: (median fluorescence intensity)
MSA: (multiple sequence alignment)
OR: (odds ratio)
PCR: (polymerase chain reaction)
PCA: (Principal component analysis)
REF: (reference)
RT: (reverse transcription)
SVD: (singular value decomposition)
SNP: (single nucleotide polymorphism)
VE: (variance explained)
MSA: (multiple sequence alignment)
RSV: (respiratory syncytial virus)

## 7 Code availability

Analysis code is available on GitHub https://github.com/DylanLawless/inspire2022lawless.github.io.

## 8 Supplemental methods

### 8.1 Study population

The study population is a longitudinal birth cohort, the INfant Susceptibility to Pulmonary Infections and Asthma Following RSV Exposure (INSPIRE), specifically designed to capture the first RSV infection in term healthy infants. Additional details of this birth cohort have been previously published [44]. Briefly, the cohort included 1949 term (≥ 37 weeks gestation), non-low birth weight (≥ 2250 g, 5 lbs), otherwise healthy infants from a population-representative sample of pediatric practices located in rural, suburban, and urban regions of the south-eastern US during 2012-2014. Infants were born June through December; per study design, they were 6 months of age or less entering their first RSV season.

### 8.2 Biweekly surveillance of RSV infection

Infant (i.e., first year of life) RSV infection was ascertained through passive and active biweekly surveillance during each infant’s first RSV season and RSV serology (Table 1). If an infant met pre-specified criteria for an acute respiratory infection, we conducted an in-person respiratory illness visit at which time we administered a parental questionnaire, performed a physical exam, collected a nasal wash, and completed a structured medical chart review for infants seen during an unscheduled visit. RSV RNA in nasal samples was detected by reverse-transcription quantitative PCR [44]. All samples were analyzed together and raw values were compared without normalization. We *a priori* defined the clinical entity of “prolonged” infection during infancy as repeatedly meeting pre-specified criteria for an acute respiratory infection accompanied by repeatedly positive RSV PCR separated by 15 or more days (Figure S1) [12].

### 8.3 Descriptive analyses

Descriptive analyses of the cohort were conducted using R 4.0.5. Pearson or Wilcoxon tests were used for comparing infants with and without prolonged RSV infection. The main descriptive features are provided in Table 1.

### 8.4 Host DNA collection and genotyping

One-year blood samples were selected based on availability of DNA among a subset of children with RSV infection and a random group of those without infection, and were genotyped with the Multi-Ethnic Global Array microarray (Illumina, CA, United States) at the University of Washington DNA Sequencing and Gene Analysis Center (Seattle, WA, United States).

### 8.5 Host genetic analyses of RSV infection in infancy

To determine whether host genetic factors are associated with infant RSV infection risk, we examined single nucleotide polymorphisms (SNPs) previously shown to alter infant RSV infection severity or childhood asthma risk [14]–[16]. For this candidate SNP analysis, we considered childhood asthma- and RSV LRTI-associated SNPs identified in Pividori et al. [14]; Janssen et al. [15]; Pasanen et al. [16]. The first is the largest childhood asthma GWAS to date, and, to our knowledge, the latter 2 represent the most comprehensive studies of RSV LRTI-associated SNPs. To further reduce the multiple testing burden, we only analysed SNPs with MAF ≥ 0.1 in at least one of the White, Black, or Hispanic ethnicity groups. We also conducted a host GWAS to identify common variants associated with infant RSV infection, and examined narrow sense heritability to test for small cumulative effects. The GWAS was performed on 621 children with available DNA for the association between host genotype and RSV infection during infancy. Due to sample size constraints, we restricted our sub-analysis to the 54 host SNPs previously associated with RSV lower respiratory tract infection or childhood asthma [14]–[16]. We additionally evaluated the accumulation of small genetic effects that would go undetected in a GWAS by estimating the narrow sense heritability of RSV infection.

For GWAS analyses, the initial round of data quality control was performed on individual populations (self-reported as White, Black, and Hispanic) using PLINK version 1.9 [43]. Subjects with a missing genotype call rate above 5% were removed. The SNP minor allele frequency (MAF) threshold was set at > 0.01, 0.03, and 0.08 for White, Black, and Hispanic, respectively [39]. The groups were merged for a total of 1,086,830 variants and a genotyping rate of 0.78. Subject independence was assessed using KING (https://people.virginia.edu/~wc9c/KING/) to prevent spurious associations. However, no probable relatives or duplicates were detected based on pairwise identify-by-state. We compared the genetic ancestry in cases to self-reported ethnicity to check for mislabelling. Reported and estimated sex was also examined for discrepancy. A second round of quality control on the combined dataset was conducted, which removed 74 samples due to genotype missingness and 399,991 variants with a genotyping rate < 0.1. Variants were checked for departure from Hardy-Weinberg equilibrium (HWE) (P < 1e-6) to uncover features of selection, population admixture, cryptic relatedness, or genotyping error. This was only performed on controls to prevent removal of genuine genetic associations that can be associated with this measurement; 6,024 variants were removed. No variants had a MAF < 0.01 after merging. SNP positions and identifiers were compared and updated according to dbNSFP4.0a (hg19) with 289 variants removed due to a missing coordinate and SNPs identifier [38]. This resulted in an analysis-ready dataset of 680,526 variants from 621 children (509 with and 112 without RSV infection in infancy), yielding a total genotyping rate of 0.98. No genomic inflation was evident with an estimated lambda (based on median chi-squared test) equal to 1. We then used genome-wide complex trait analysis (GCTA) software (https://cnsgenomics.com/software/gcta/) to calculate the genetic relationship matrix and performed principal component analysis (PCA) to account for population structure [39].

Genome-wide association analysis was performed using PLINK version 1.9 for logistic regression with multiple covariates consisting of the child’s birth month, enrolment year (as a marker of RSV season), daycare attendance, presence of another child ≤ 6 years of age at home, sex, and 6 ancestry principal components (PCs) [43].

As the multiple testing burden likely precluded identification of small genetic effects in our GWAS, we conducted an additional heritability analysis using the method described by Golan et al. [17] to estimate narrow-sense heritability of RSV infection during infancy on the latent liability scale (h^2^_l_), which, if > 0, would indicate an accumulation of small genetic effects. We estimated h^2^_l_ to be exactly 0, suggesting that, if present, infant RSV infection-related genetic signals are both small and sparse.

### 8.6 Host genetic analyses for known associations

We further investigated the possibility that the analysis was underpowered to identify associations with reported childhood asthma- and RSV LRTI-associated SNPs [14]–[16]. This was done by pooling information across SNPs to estimate the average genetic effect size. In brief, we computed a z-score for each SNP, where the average (across SNPs) squared z-score G(bar) is proportional to the average squared genetic effect on RSV infection in infancy. As G(bar) is an average of p = 54 approximately independent statistics, it is approximately N (nμ^2^ + 1, 2/p), where n = 621 is the sample size and μ^2^ is a function of the average squared genetic effect on RSV infection in infancy. Using the genetic effect estimates from Pividori et al. [14]; Janssen et al. [15]; Pasanen et al. [16], we calculated that we would have 80% power to reject the global null hypothesis of no genetic effect at any of these SNPs (i.e., μ^2^ = 0) if, on average across the 54 SNPs, the genetic effect on RSV infection in infancy was at least 61% as large as those estimated in the aforementioned 3 studies. We found G(bar)=1.00 in our data, which corresponds to a p value of 0.50. This result indicates that the genetic effect on RSV infection in infancy is zero or small at SNPs likely to be associated with RSV infection *a priori*.

### 8.7 Host acute local immune response

Nasal wash samples collected at the time of acute infant RSV infection were profiled to measure the acute host response to infection using Luminex xMap multianalyte bead assays (Milliplex Human Cytokine/Chemokine Panel II MAGNETIC Premixed 23 Plex Kit, EMD Millipore; and Cytokine 30-Plex Human Panel, Life Technologies Corporation). These data were used to test the host nasal interferon (IFN) response and the viral variant associated with prolonged infection.

### 8.8 RSV whole-genome sequencing

RSV genome sequencing was performed on all specimens from subjects meeting illness criteria and with positive RSV PCR. Viral amino acid variants (genotype) of the F and G glycoprotein were tested for association with prolonged infection adjusting for host features associated with increased infection risk. The relatively small sample size of our cohort required analysis that targeted only genes which were a priori likely to functionally contribute to the clinical phenotype. Therefore, our analysis focused on the surface F (fusion) and G (attachment) proteins of RSV as they have been implicated in pathogenesis [45], [46], and both are targets for neutralizing antibodies during infection [47], [48]. Lastly, to determine if the variants of interest were enriched by selective pressure over time, we used public data from the past several decades to assess variant frequency over time.

RSV whole-genome sequencing of this study population has been previously described [49]. Briefly, RNA was extracted at J. Craig Venter Institute (JCVI) (https://www.jcvi.org) in Rockville, MD from nasal wash samples which were RSV PCR positive and collected during a respiratory illness visit triggered through biweekly surveillance of symptoms. Four forward reverse-transcription (RT) primers were designed and four sets of PCR primers were manually picked across a consensus of complete RSV genome sequences using JCVI’s automated primer design tool [50]. cDNA was generated from 4 μL undiluted RNA using the pooled forward primers and SuperScript III Reverse Transcriptase (Thermo Fisher Scientific, Waltham, MA, USA). 100 ng of pooled DNA amplicons were sheared to create 400-bp libraries, which were pooled in equal volumes and cleaned with Ampure XP reagent (Beckman Coulter, Inc., Brea, CA, USA). Sequencing was performed on the Ion Torrent PGM using 316v2 or 318v2 chips (Thermo Fisher Scientific).

For samples requiring extra coverage, in addition to Ion Torrent sequencing, Illumina libraries were prepared using the Nextera DNA Sample Preparation Kit (Illumina, Inc., San Diego, CA, USA). Sequence reads were sorted by barcode, trimmed, and assembled de novo using CLC Bio’s *clc novo assemble* program, and the resulting contigs were searched against custom, full-length RSV nucleotide databases to find the closest reference sequence. All sequence reads were then mapped to the selected reference RSV sequence using CLC Bio’s *clc ref assemble long* program [51]. Curated assemblies were validated and annotated with the viral annotation software called Viral Genome ORF Reader, VIGOR 3.0 (https://sourceforge.net/projects/jcvi-vigor/files/), before submission to GenBank as part of the Bioproject accession PRJNA225816 (https://www.ncbi.nlm.nih.gov/bioproject/225816) [52] and PRJNA267583 (https://www.ncbi.nlm.nih.gov/bioproject/267583).

### 8.9 Viral sequence alignment

The NCBI-tools, Tbl2asn (https://www.ncbi.nlm.nih.gov/genbank/tbl2asn2/), was used in the creation of sequence records for submission to GenBank (https://www.ncbi.nlm.nih.gov/genbank/). A total of 350 viral sequences in .sqn file format were used for downstream analysis.

We computed a phylogenetic tree for each gene, as follows. NCBI-tools asn2fsa (https://www.huge-man-linux.net/man1/asn2fsa.html) was used to convert sequences to fasta format. Each sample consisted of 11 sequence segments (NS1, NS2, N, P, M, M2-1, M2-2, SH, G, F, and L) as shown in Figure 1. These were separated and repooled to create 11 single fasta files for each gene containing all 350 samples. Sequences were checked for at least 90% coverage of the corresponding gene to minimize loss of aligned positions when computing the phylogenetic tree. Each of the eleven resulting sets was aligned with MAFFT v7 (https://mafft.cbrc.jp/alignment/software/) [42], using default parameters. The sequence of the orthologous gene from Bovine orthopneumovirus (GenBank:NC 001989) was added to each set as an outgroup.

IQ-Tree (https://www.iqtree.org) [40] was used with per-gene multiple sequence alignment (MSA) files based on amino acid sequence for estimating maximum-likelihood phylogenies using protein substitution model. Examining the sequences with an alignment viewer showed that a small number of sequences had frame-shift variants but these did not affect the regions included in our testing criteria.

Viral sequence data and clinical information were merged and cleaned with R. Clinical IDs matching more than one viral sequence ID were used to re-identify samples from the same individual as prolonged infections. Genetic variation was quantified in these samples, and for subsequent analysis, only the first viral sequence was included for association testing. Antigenic grouping of strain A and B had been completed previously and labels were included to annotate each sample accordingly.

The cohort-specific variant frequency per position was calculated; residues were counted and ranked by frequency with the most frequent residue defined as reference (REF) and alternative (ALT) for variants. Positions with at least one ALT were checked for potential misalignment or other sources of error. Variant positions were selected for association analysis, while non-variant positions were ignored.

A number of host features have been previously shown to influence infection susceptibility and were therefore included as covariates in our analysis [53]. Six samples were excluded due to insufficient covariate data, resulting in 344 test samples. Of these, 38 were from the same patients (prolonged infection) of which half (19) were included for association testing. Thus, the test set was comprised of single samples collected from 325 individuals.

### 8.10 Viral population structure

The genetic distances to nearest neighbors were computed based on phylogenetic trees generated with MAFFT. PCA and singular value decomposition (SVD) were used in dimensionality reduction for exploratory data analysis of viral phylogeny. The R package factoextra was used for PCA and to visualise eigenvalues and variance. R package caret was used to analyse genetic correlations.

The duration of RSV shedding in Kenyan infants has been reported previously [12]. Based on these findings, infection events separated by at least 15 days with symptoms were expected to be “new” infections [12]. To determine whether infections were new or prolonged (i.e., failure to clear), the pairwise genetic distance was compared between every viral sequence. Small distances indicate pairs with closely related sequences, while no detectable variation indicated a prolonged infection.

### 8.11 Viral variant association testing

The consensus sequence within the cohort was assigned based on the major allele. Variants at the amino acid level were defined as either reference (REF) or alternative (ALT). Viral amino acids (genotype collapsed into REF/ALT) were tested for association with infection types (i.e., resolved and prolonged) including key covariates that alter infection risk. To reduce the multiple testing burden, proxy amino acid variants were identified by performing clumping with ranking based on MAF and with a cut-off threshold of r^2^ ≥ 0.8 (Supplemental Figure S3). Since many variants within RSV coding genes have non-random association due to selection, analogous to linkage disequilibrium (LD) in human GWAS, we reduced the multiple testing burden by retaining proxy variants and removing those with r^2^ ≥ 0.8. Analysis was performed using logistic regression with the R stats (3.6.2) glm function. The model consisted of the binary response (prolonged infection Yes/No) and predictors viral genotype (REF/ALT amino acid, including multi-allelic non-REF collapsed into ALT), viral PCs 1-5, host sex, and host features that have been previously demonstrated as significantly associated with infection: daycare attendance, living with siblings and self-reported race/ethnicity [53].

Environmental host covariates did not contribute significant effect in our model for candidate causal association. For this reason, in our main analysis, viral population structure was accounted for by the first five PCs. The Bonferroni correction for multiple testing was applied based on the number of variants tested. For the significant association found by proxy amino acid variants, the association test was repeated for all clumped variants to produce a LocusZoom-style Manhattan plot containing r^2^ by color and p value statistics. R package stats was used for a range of analyses including glm for logistic regressions. R package MASS was used to analyse logistic regression model data. To test if the significantly associated variants were due to population structure, we re-estimated models using the subset of individuals infected with RSV strain B to confirm validity of combined analysis.

### 8.12 Public viral sequence data

We gathered publicly available sequence data to further assess variants of interest (p.E123K/D, p.P218T/S/L, and any non-specific gene-level event). We used the public viral data repository of NCBI https://www.ncbi.nlm.nih.gov/labs/virus/vssi/#/virus?SeqType_s=Nucleotide&VirusLineage_ss=Human%20orthopneumovirus,%20taxid:11250 to retrieve information using search criteria that follow. Virus: Human orthopneumovirus (HRSV), taxid:11250. Proteins: attachment glycoprotein. Host: Homo (humans), taxid:9605. Collection dates: Jan 1, 1956 onward. Nucleotide and protein sequence data was collected, which contained data from 27 countries and 1084 glycoprotein protein sequences after curation. Sequence and meta data were merged. Multiple sequence alignment was performed to find consensus relative positions for all sequences. Regions of interest were then extracted and re-annotated with their correct amino acid positions matching the reference sequence. Summary statistics were generated, including number of samples, collection date, geo-location, variant frequency, and strain for the specified amino acid (Supplemental Figure S4).

### 8.13 Biological interpretation

As infant RSV infection stimulates an acute antiviral response and also results in decreased barrier function of the airway epithelium [37], we tested for association between host interferon (IFN) response and the amino acid (REF/ALT) identified as the viral variant associated with prolonged infection. A Wilcoxon test was performed to compare IFN-γ and IFN-α between RSV amino acid positions, with adjustment for the same covariates as in the main analysis. Protein structures were analysed with data sourced from RCSB PDB https://www.rcsb.org. Protein function and domains were assessed using UniProt (https://www.uniprot.org) for P03423 (GLYC HRSVA) (strain A2) and O36633 (GLYC HRSVB) (strain B1) in gff format (https://www.uniprot.org/uniprot/P03423 and https://www.uniprot.org/uniprot/O36633, respectively). Interactions, post-translational modifications, motifs, and epitopes were assessed from the literature. Protein features were assessed using data from NCBI (https://www.ncbi.nlm.nih.gov/ipg/NP_056862.1) and via sequence viewer with O36633.1 human RSV B1 (https://www.ncbi.nlm.nih.gov/projects/sviewer/?id=O36633.1) Potential effects of amino acid variation on protein structure and function were considered according to available information on a broad range of biological and biochemical features, including native conformation (secondary, tertiary, and quaternary), domains and topology, disulfide bonds, glycosylation, interactions with other viral proteins and host-cell factors, proteolytic cleavage sites, normal patterns of intra-and/or extra-cellular distribution, and secretion status.

## 9 Supplemental results

### 9.1 Host response

Prolonged infections associated with G protein variants p.E123K/D and p.P218T/S/L were on average less severe compared with other circulating variants, and all were limited to the upper respiratory tract (Table 1). Therefore, we analysed nasal wash samples collected during acute RSV infection for a panel of cytokines involved in antiviral immune responses and observed differential IFNα and IFNγ levels segregating according to viral antigenic group—A or B. Both cytokines were elevated in group B infections compared to group A. The groups A and B median (lower-and upper-quartile) values were 9.5 (3-22.5) and 12.6 (4.1-25.8) median fluorescence intensity (MFI) units, respectively, for IFNα and 3.6 (1-7) and 4 (2-7.4) MFI, units respectively, for IFNγ (group A, n = 149; group B, n = 103). As prolonged infections with p.E123K/D and p.P218T/S/L genotypes were exclusively group B, the dichotomous relationship of IFNα and IFNγ levels to antigenic group precluded evaluation of G protein variants as independent predictors of IFNα and IFNγ production.

Continued form main text

## Supplemental figures

**Figure S1:**
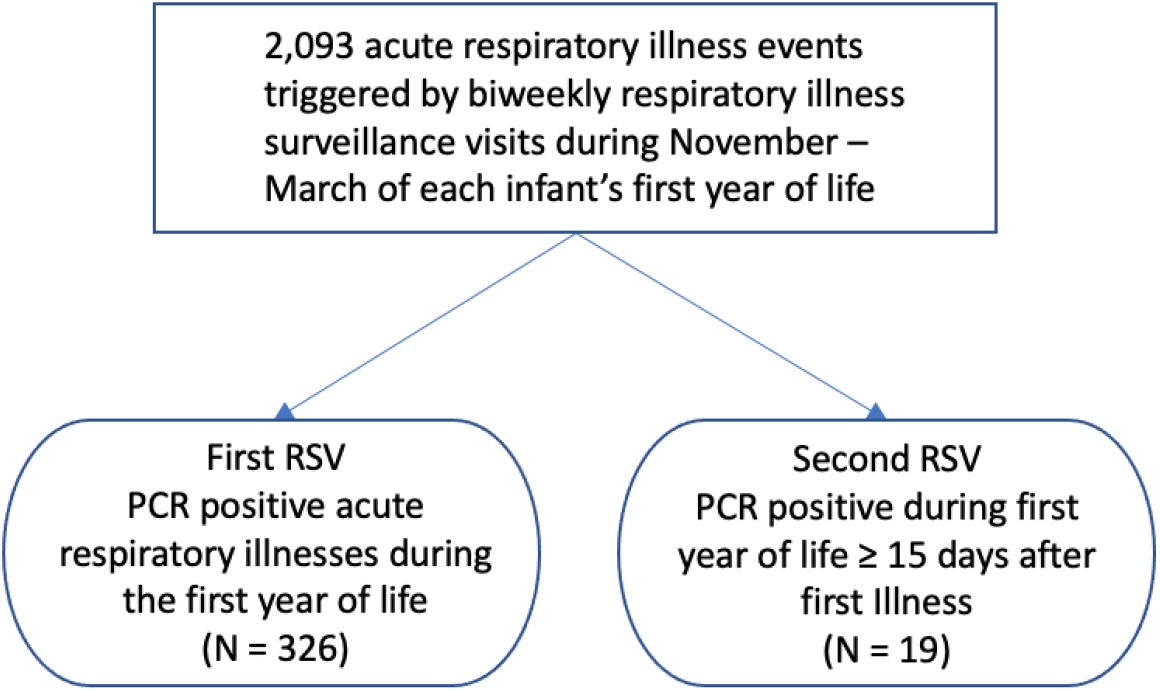
INfant Susceptibility to Pulmonary Infections and Asthma Following RSV Exposure (INSPIRE). The study population is a longitudinal birth cohort specifically designed to capture the first RSV infection in term healthy infants. Prolonged infection was a priori defined as repeatedly meeting criteria for acute respiratory infection with RSV PCR positive nasal samples ≥ 15 days between testing.

**Figure S2:**
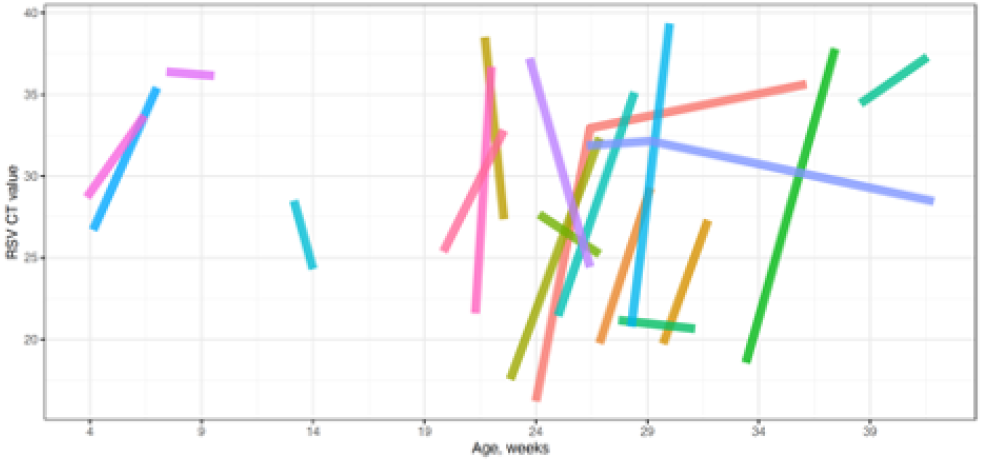
Supplemental: Infant RSV prolonged infections. Each line represents an infant in the study, and line start and end correspond to clinical respiratory illness sampling timepoints. CT values are inversely related to viral RNA abundance.

**Figure S3:**
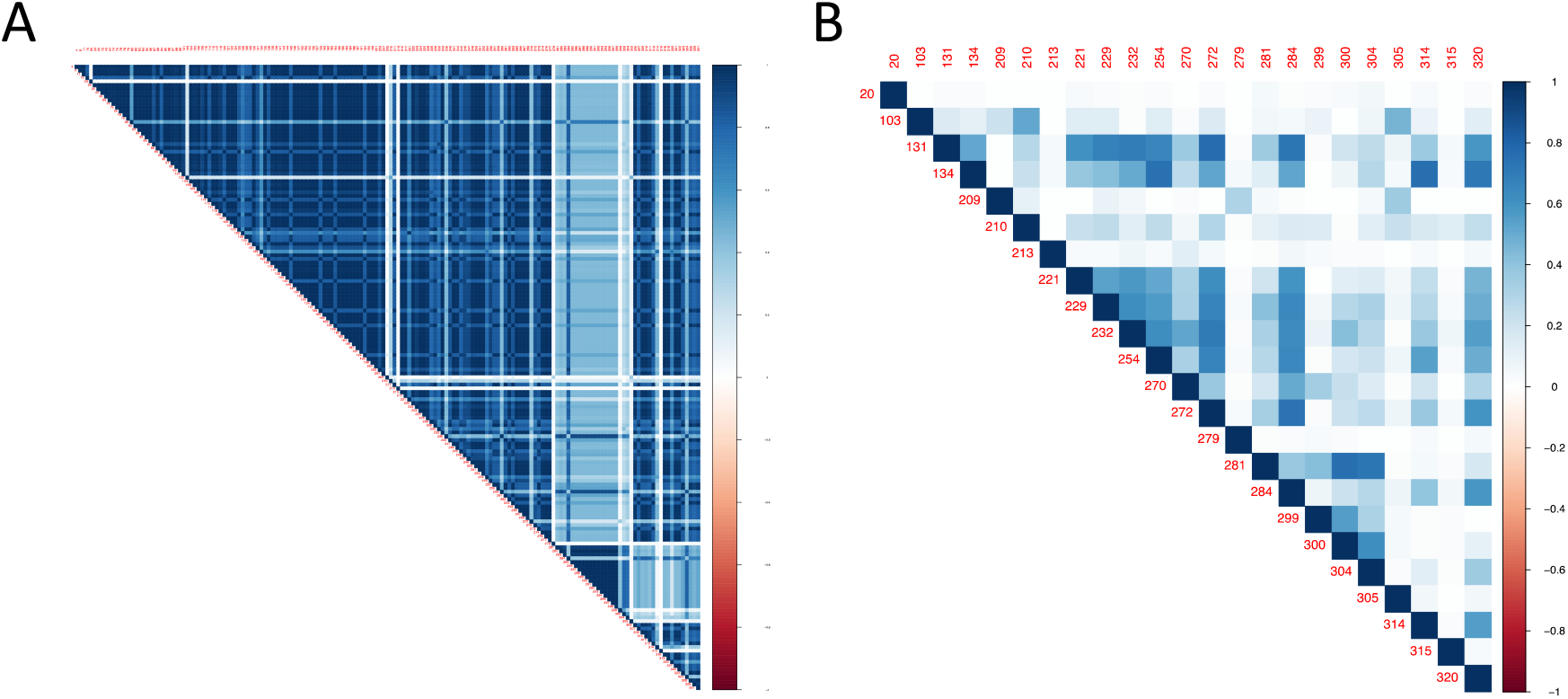
Supplemental: Variant clumping for reduction in association testing. [Left] Correlation between all positions. [Right] Correlation between proxy variants after clumping to remove r2 ≥ 0.8. Values indicate relative amino acid positions within MSA. r2 indicated by color scale.

**Figure S4:**
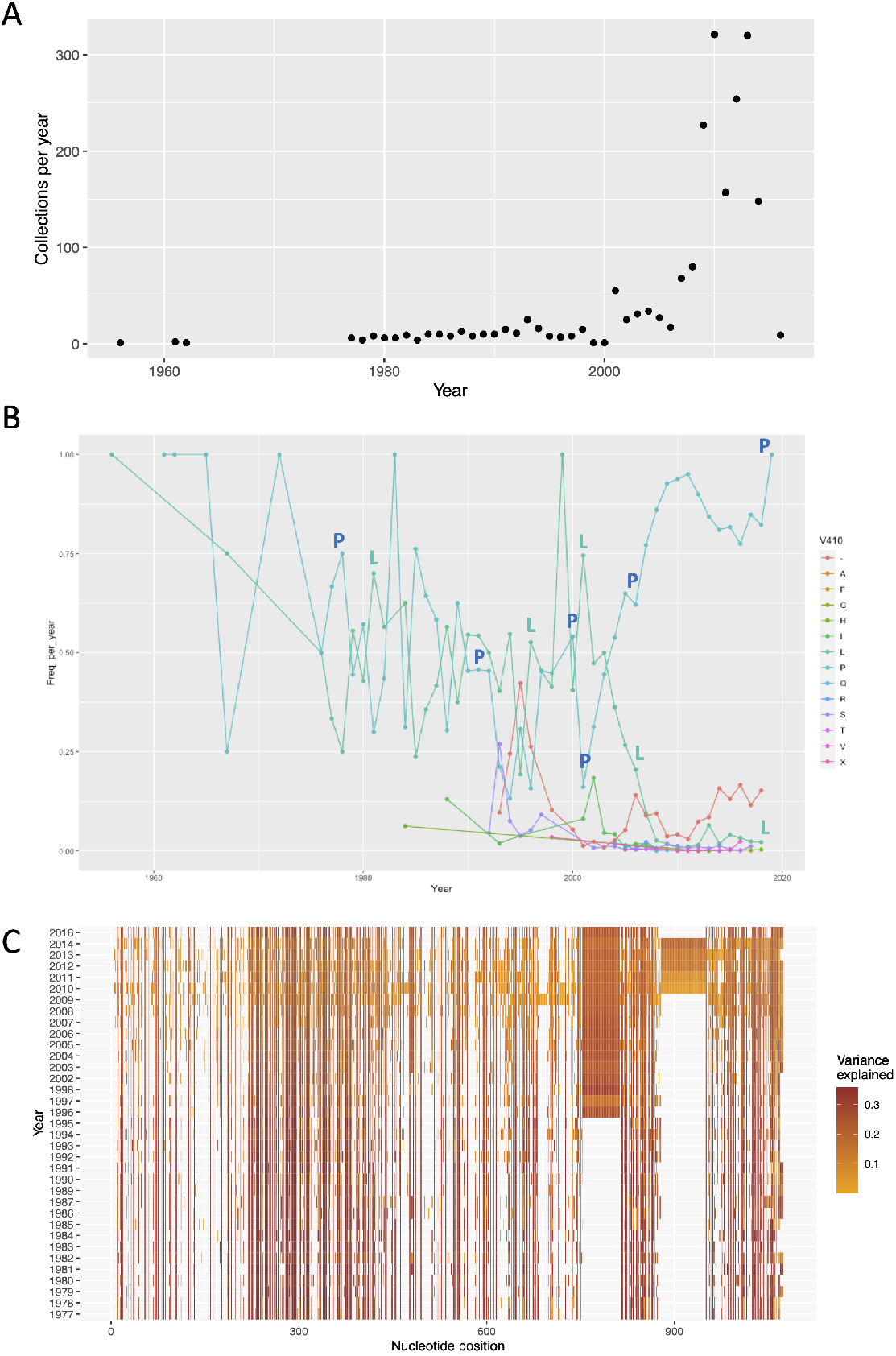
Supplemental: Publicly available RSV sequence data for > 30 years. (A) Global sample collection per year. (B) Variant associated with prolonged infection tracked in public data. The lead proxy SNP, p.P218T/S/L is illustrated here (relative amino acid positive 410 in MSA). The major alleles (proline, leucine) are seen for group A/B, with minor alleles (serine, threonine) generally at low frequency <10%. (C) % variance explained per year for all G protein amino acid variants from 1990-2022.

## References

[1] C. B. Hall et al., “The burden of respiratory syncytial virus infection in young children,” N Engl J Med, vol. 360, no. 6, pp. 588–98, Feb. 2009, doi: 10.1056/NEJMoa0804877.

[2] W. P. Glezen, L. H. Taber, A. L. Frank, and J. A. Kasel, “Risk of primary infection and reinfection with respiratory syncytial virus,” Am J Dis Child, vol. 140, no. 6, pp. 543–6, Jun. 1986, doi: 10.1001/archpedi.1986.02140200053026.

[3] V. Bokun et al., “Respiratory syncytial virus exhibits differential tropism for distinct human placental cell types with Hofbauer cells acting as a permissive reservoir for infection,” PLoS One, vol. 14, no. 12, p. e0225767, 2019, doi: 10.1371/journal.pone.0225767.

[4] H. A. Cubie, L. A. Duncan, L. A. Marshall, and N. M. Smith, “Detection of respiratory syncytial virus nucleic acid in archival postmortem tissue from infants,” Pediatr Pathol Lab Med, vol. 17, no. 6, pp. 927–38, Nov. 1997.

[5] D. Nadal, W. Wunderli, O. Meurmann, J. Briner, and J. Hirsig, “Isolation of respiratory syncytial virus from liver tissue and extrahepatic biliary atresia material,” Scand J Infect Dis, vol. 22, no. 1, pp. 91–3, 1990, doi: 10.3109/00365549009023125.

[6] D. R. O’Donnell, M. J. McGarvey, J. M. Tully, I. M. Balfour-Lynn, and P. J. Openshaw, “Respiratory syncytial virus RNA in cells from the peripheral blood during acute infection,” J Pediatr, vol. 133, no. 2, pp. 272–4, Aug. 1998, doi: 10.1016/s0022-3476(98)70234-3.

[7] F. Rezaee, L. F. Gibson, D. Piktel, S. Othumpangat, and G. Piedimonte, “Respiratory syncytial virus infection in human bone marrow stromal cells,” Am J Respir Cell Mol Biol, vol. 45, no. 2, pp. 277–86, Aug. 2011, doi: 10.1165/rcmb.2010-0121OC.

[8] A. Rohwedder, O. Keminer, J. Forster, K. Schneider, E. Schneider, and H. Werchau, “Detection of respiratory syncytial virus RNA in blood of neonates by polymerase chain reaction,” J Med Virol, vol. 54, no. 4, pp. 320–7, Apr. 1998, doi: 10.1002/(sici)1096-9071(199804)54:4<320::aid-jmv13>3.0.co;2-j.

[9] R. E. Randall and D. E. Griffin, “Within host RNA virus persistence: mechanisms and consequences,” Current opinion in virology, vol. 23, pp. 35–42, 2017.

[10] P. K. Munywoki et al., “Influence of age, severity of infection, and co-infection on the duration of respiratory syncytial virus (RSV) shedding,” Epidemiol Infect, vol. 143, no. 4, pp. 804–12, Mar. 2015, doi: 10.1017/s0950268814001393.

[11] B. Bagga, L. Harrison, P. Roddam, and J. DeVincenzo, “Unrecognized prolonged viral replication in the pathogenesis of human RSV infection,” Journal of Clinical Virology, vol. 106, pp. 1–6, 2018.

[12] E. A. Okiro, L. J. White, M. Ngama, P. A. Cane, G. F. Medley, and D. J. Nokes, “Duration of shedding of respiratory syncytial virus in a community study of Kenyan children,” BMC infectious diseases, vol. 10, no. 1, pp. 1–7, 2010.

[13] E. K. Larkin and T. V. Hartert, “Genes associated with RSV lower respiratory tract infection and asthma: the application of genetic epidemiological methods to understand causality,” Future virology, vol. 10, no. 7, pp. 883–897, 2015.

[14] M. Pividori, N. Schoettler, D. L. Nicolae, C. Ober, and H. K. Im, “Shared and distinct genetic risk factors for childhood-onset and adult-onset asthma: genome-wide and transcriptome-wide studies,” The Lancet Respiratory Medicine, vol. 7, no. 6, pp. 509–522, 2019.

[15] R. Janssen et al., “Genetic susceptibility to respiratory syncytial virus bronchiolitis is predominantly associated with innate immune genes,” Journal of Infectious Diseases, vol. 196, no. 6, pp. 826–834, 2007.

[16] A. Pasanen et al., “Genome-wide association study of polymorphisms predisposing to bronchiolitis,” Scientific reports, vol. 7, no. 1, pp. 1–9, 2017.

[17] D. Golan, E. S. Lander, and S. Rosset, “Measuring missing heritability: inferring the contribution of common variants,” Proceedings of the National Academy of Sciences, vol. 111, no. 49, pp. E5272–E5281, 2014.

[18] A. Eshaghi et al., “Genetic variability of human respiratory syncytial virus A strains circulating in Ontario: a novel genotype with a 72 nucleotide G gene duplication,” PloS one, vol. 7, no. 3, p. e32807, 2012.

[19] C. B. Hall, J. M. Geiman, R. Biggar, D. I. Kotok, P. M. Hogan, and R. G. Douglas Jr, “Respiratory syncytial virus infections within families,” New England journal of medicine, vol. 294, no. 8, pp. 414–419, 1976.

[20] S. Levine, R. Klaiber-Franco, and P. Paradiso, “Demonstration that glycoprotein G is the attachment protein of respiratory syncytial virus,” Journal of General Virology, vol. 68, no. 9, pp. 2521–2524, 1987.

[21] S. A. Feldman, R. M. Hendry, and J. A. Beeler, “Identification of a linear heparin binding domain for human respiratory syncytial virus attachment glycoprotein G,” Journal of virology, vol. 73, no. 8, pp. 6610–6617, 1999.

[22] S. A. Feldman, S. Audet, and J. A. Beeler, “The fusion glycoprotein of human respiratory syncytial virus facilitates virus attachment and infectivity via an interaction with cellular heparan sulfate,” Journal of Virology, vol. 74, no. 14, pp. 6442–6447, 2000.

[23] H. M. Rixon, G. Brown, J. Murray, and R. Sugrue, “The respiratory syncytial virus small hydrophobic protein is phosphorylated via a mitogen-activated protein kinase p38-dependent tyrosine kinase activity during virus infection,” Journal of General Virology, vol. 86, no. 2, pp. 375–384, 2005.

[24] R. Ghildyal et al., “Interaction between the respiratory syncytial virus G glycoprotein cytoplasmic domain and the matrix protein,” Journal of General Virology, vol. 86, no. 7, pp. 1879–1884, 2005.

[25] P. L. Collins and G. Mottet, “Oligomerization and post-translational processing of glycoprotein G of human respiratory syncytial virus: altered O-glycosylation in the presence of brefeldin A,” Journal of General Virology, vol. 73, no. 4, pp. 849–863, 1992.

[26] M. E. Monzon, S. M. Casalino-Matsuda, and R. M. Forteza, “Identification of glycosaminoglycans in human airway secretions.,” Am J Respir Cell Mol Biol, vol. 34, no. 2, pp. 135–141, Feb. 2006, doi: 10.1165/rcmb.2005-0256OC.

[27] S. M. Johnson et al., “Respiratory syncytial virus uses CX3CR1 as a receptor on primary human airway epithelial cultures,” PLoS pathogens, vol. 11, no. 12, p. e1005318, 2015.

[28] H.-J. Lee, J.-Y. Lee, M.-H. Park, J.-Y. Kim, and J. Chang, “Monoclonal Antibody against G Glycoprotein Increases Respiratory Syncytial Virus Clearance In Vivo and Prevents Vaccine-Enhanced Diseases.,” PLoS One, vol. 12, no. 1, p. e0169139, 2017, doi: 10.1371/journal.pone.0169139.

[29] E. J. Collarini et al., “Potent High-Affinity Antibodies for Treatment and Prophylaxis of Respiratory Syncytial Virus Derived from B Cells of Infected Patients,” J. Immunol., vol. 183, no. 10, p. 6338, Nov. 2009, doi: 10.4049/jimmunol.0901373.

[30] M. E. Brint et al., “Prolonged viral replication and longitudinal viral dynamic differences among respiratory syncytial virus infected infants,” Pediatric research, vol. 82, no. 5, pp. 872–880, 2017.

[31] R. A. Tripp, L. P. Jones, L. M. Haynes, H. Zheng, P. M. Murphy, and L. J. Anderson, “CX3C chemokine mimicry by respiratory syncytial virus G glycoprotein,” Nature immunology, vol. 2, no. 8, pp. 732–738, 2001.

[32] K.-I. Jeong et al., “CX3CR1 is expressed in differentiated human ciliated airway cells and co-localizes with respiratory syncytial virus on cilia in a G protein-dependent manner,” PloS one, vol. 10, no. 6, p. e0130517, 2015.

[33] L. Zhang et al., “Infection of ciliated cells by human parainfluenza virus type 3 in an in vitro model of human airway epithelium.,” J Virol, vol. 79, no. 2, pp. 1113–1124, Jan. 2005, doi: 10.1128/JVI.79.2.1113-1124.2005.

[34] A. Bukreyev et al., “The secreted form of respiratory syncytial virus G glycoprotein helps the virus evade antibody-mediated restriction of replication by acting as an antigen decoy and through effects on Fc receptor-bearing leukocytes,” Journal of virology, vol. 82, no. 24, pp. 12191–12204, 2008.

[35] M. E. Schmidt and S. M. Varga, “Modulation of the host immune response by respiratory syncytial virus proteins,” Journal of Microbiology, vol. 55, no. 3, pp. 161–171, 2017.

[36] T. Chirkova et al., “Effect of Infant RSV Infection on Memory T Cell Responses at Age 2-3 Years,” Frontiers in immunology, vol. 13, p. 826666, 2022.

[37] A. R. Connelly et al., “Metabolic Reprogramming of Nasal Airway Epithelial Cells Following Infant Respiratory Syncytial Virus Infection,” Viruses, vol. 13, no. 10, p. 2055, 2021.

[38] X. Liu, C. Wu, C. Li, and E. Boerwinkle, “dbNSFP v3.0: A One-Stop Database of Functional Predictions and Annotations for Human Nonsynonymous and Splice-Site SNVs,” Human Mutation, vol. 37, no. 3, pp. 235–241, 2016, doi: 10.1002/humu.22932.

[39] J. Yang, S. H. Lee, M. E. Goddard, and P. M. Visscher, “GCTA: a tool for genome-wide complex trait analysis,” The American Journal of Human Genetics, vol. 88, no. 1, pp. 76–82, 2011.

[40] L.-T. Nguyen, H. A. Schmidt, A. Von Haeseler, and B. Q. Minh, “IQ-TREE: a fast and effective stochastic algorithm for estimating maximum-likelihood phylogenies,” Molecular biology and evolution, vol. 32, no. 1, pp. 268–274, 2015.

[41] A. Manichaikul, J. C. Mychaleckyj, S. S. Rich, K. Daly, M. Sale, and W.-M. Chen, “Robust relationship inference in genome-wide association studies,” Bioinformatics, vol. 26, no. 22, pp. 2867–2873, 2010.

[42] K. Katoh and D. M. Standley, “MAFFT multiple sequence alignment software version 7: improvements in performance and usability,” Molecular biology and evolution, vol. 30, no. 4, pp. 772–780, 2013.

[43] S. Purcell et al., “PLINK: a tool set for whole-genome association and population-based linkage analyses,” The American journal of human genetics, vol. 81, no. 3, pp. 559–575, 2007.

[44] E. K. Larkin et al., “Objectives, design and enrollment results from the Infant Susceptibility to Pulmonary Infections and Asthma Following RSV Exposure Study (INSPIRE),” BMC Pulm Med, vol. 15, p. 45, Apr. 2015, doi: 10.1186/s12890-015-0040-0.

[45] S. Boyoglu-Barnum et al., “An anti-G protein monoclonal antibody treats RSV disease more effectively than an anti-F monoclonal antibody in BALB/c mice,” Virology, vol. 483, pp. 117–125, 2015.

[46] A. Bukreyev, L. Yang, and P. L. Collins, “The secreted G protein of human respiratory syncytial virus antagonizes antibody-mediated restriction of replication involving macrophages and complement,” Journal of virology, vol. 86, no. 19, pp. 10880–10884, 2012.

[47] L. J. Anderson, P. Bingham, and J. Hierholzer, “Neutralization of respiratory syncytial virus by individual and mixtures of F and G protein monoclonal antibodies,” Journal of virology, vol. 62, no. 11, pp. 4232–4238, 1988.

[48] J. O. Ngwuta et al., “Prefusion F–specific antibodies determine the magnitude of RSV neutralizing activity in human sera,” Science translational medicine, vol. 7, no. 309, pp. 309ra162–309ra162, 2015.

[49] S. A. Schobel et al., “Respiratory Syncytial Virus whole-genome sequencing identifies convergent evolution of sequence duplication in the C-terminus of the G gene,” Sci Rep, vol. 6, p. 26311, May 2016, doi: 10.1038/srep26311.

[50] K. Li et al., “Automated degenerate PCR primer design for high-throughput sequencing improves efficiency of viral sequencing,” Virol J, vol. 9, p. 261, Nov. 2012, doi: 10.1186/1743-422x-9-261.

[51] Q. Aarhus, “White paper on de novo assembly in CLC Assembly Cell 4.0,” digitalinsights, p. 14, Jun. 2016.

[52] S. Wang, J. P. Sundaram, and T. B. Stockwell, “VIGOR extended to annotate genomes for additional 12 different viruses,” Nucleic Acids Res, vol. 40, no. Web Server issue, pp. W186–92, Jul. 2012, doi: 10.1093/nar/gks528.

[53] C. Rosas-Salazar et al., “Upper respiratory tract bacterial-immune interactions during respiratory syncytial virus infection in infancy,” Journal of Allergy and Clinical Immunology, vol. 149, no. 3, pp. 966–976, 2022.

